# Prevalence and factors associated with electrocardiographic abnormalities among adults attending Methadone Assisted Therapy in Dodoma, central Tanzania

**DOI:** 10.1101/2022.11.21.22282602

**Authors:** Immaculate Kalungi, Martin Mujuni, Innocent Mwombeki, Azan Nyundo

## Abstract

**Background:** Patients with opioid use disorder are at a higher cardiovascular risk due to the effect of opioids on the cardiovascular system. Cardiac conduction abnormalities, electrical activity impairment, cardiac arrhythmias, and ventricular hypertrophy are reported in the opioid population.

**Objective:** This study aimed to assess the prevalence and factors associated with ECG abnormalities among adults with opioid use disorder attending the Itega addiction center for methadone-assisted therapy (MAT).

**Methodology:** A cross-sectional analytical study was conducted among adult outpatients attending the Itega addiction center in Dodoma. A calculated sample size of 321was attained through a convenience sampling approach. A standard 12-lead ECG was recorded for each participant and interpreted by two independent cardiologists. Univariate and multivariable logistic regression was computed to determine the factors associated with ECG abnormalities. Under adjusted analysis, a p-value of less than 0.05 was considered significant for factors associated with ECG abnormalities after controlling for all the variables with a minimum p-value of 0.2 at univariate analysis.

**Results:** The majority of 308 (95.95%) of the participants were males, 197 (61.37%) had attained primary education level, and the mean age of the participants was 35.44 ± 6.54 years. The overall prevalence for any ECG abnormalities in this study was 26.47%, with Sinus bradycardia 59(18.4%) being the most observed ECG abnormality, followed by QTc prolongation 27(8.41%). A month’s increase in the duration on MAT and being a female were significantly associated with lower odds of ECG abnormalities (AOR =0.85, 95% CI =0.74-0.96 p =0.014) and (AOR = 0.05, 95% CI = 0.01-0.59, p = 0.017) respectively.

**Conclusion:** The high prevalence of ECG Abnormalities implies high cardiovascular risk among a population with opioid use disorder. Given that majority of the ECG abnormalities are treatable, integrating cardiovascular care in the opioid addiction clinic would be beneficial for this population.

## INTRODUCTION

Opioid use continues to be a significant problem of public health importance worldwide, as per the 2019 report; 53 million people are estimated to use opioids, and opioids were responsible for two-thirds of 587,000 drug-related death by 2017(1). At the same time, up to 300,000 people are estimated to use opioids, with significant proportion affected by HIV/AIDS and Hepatitis B virus(2). Heroin is particularly the most abused opioid worldwide, in the United States; 808,000 (0.3%) of people aged 12 years and older used heroin between 2017-2018, while 157,000(0.5%) of the young adult populations aged 18-25 years use heroin in the same period(3).

Although the a biological pathway between opioid exposure and cardiovascular disease(CVD), there is an association between opioid drug use and increased risk of a cardiovascular event such as heart failure (MI) and myocardial infarction(HF) (4). Electrocardiographic abnormalities in the opioid population; include arrhythmias, sinus bradycardia, hypotension, and benign atrial-ventricular block. Also, the resulting atrial or ventricular automatic ectopy and tachycardia(5). Several factors have been associated with ECG abnormalities in opioid use, including sex, age, comorbid diseases prior to treatment, duration on methadone treatment, other prescription drugs which interact with methadone, and concomitant use of other illicit drugs while on treatment(6).

While there are various ECG abnormalities among patients with opioid use disorders, most studies have focused mainly on QTc prolongation. Furthermore, no studies in Tanzania report the burden and predictors of cardiovascular abnormalities in the opioid population. This study, therefore, aims to determine the prevalence and factors associated with ECG abnormalities among adults with opioid use disorders attending Methadone Assisted Therapy at the Itega addiction centre in Dodoma, Tanzania.

## Materials and Methods

Study design

A cross-sectional analytical study

### Study site and population

The study was carried out at Itega MAT clinic located at Itega addiction centre, a satellite unit of Mirembe mental health hospital (The National Mental Hospital) in Dodoma city, Tanzania, with 500 enrolled clients at the time of the study. Itega addiction clinic is located in the Itega area, and has one psychiatrist, five medical officers, four nurses, one social worker, one occupational therapist, one laboratory scientist, and two pharmacists. The clinic is conducted every day from 7.30hrs to 15.30hrs, attending up to 300 patients per day with opioid and other drug use disorders. Before initiation of treatment, all patients are counseled and screened for drug use, comorbid psychiatric conditions, HIV, and Hepatitis B and C. The patients attend daily for the methadone dose, which is given as a solution. The initial starting dose is 30 mg, increased gradually according to the patient’s symptoms of withdrawal.

### Sample size

A sample size of 316 was calculated using the Kish and Leslie formula, N = Z^2^ (P (1-P)/d^2^), where: N = sample size, Z=Score for 95% Confidence interval, which is 1.96, P=Prevalence of ECG abnormalities in Methadone assisted clinic in a previous study (7), p=29%, d=Marginal error set at 5%. With a 5% contingency, 16 participants were added to make a final sample size of 332.

### Sampling techniques

Non-probability sampling (consecutive sampling) was applied whereby all clients attending at Itega MAT clinic who met the inclusion criteria were recruited in the study. The participants were enrolled consecutively, whereby the first contact with the participants was at the waiting lounge before going to the clinician for daily care after routine registration. They were then introduced to the overall procedures of the study and asked to participate after receiving their routine daily care. Study procedures were applied consecutively from one client to the next without skipping until the sample size was reached.

### Inclusion and exclusion criteria

The study included all patients above 18 years attending the Itega MAT clinic who can provide informed consent and must be on steady methadone dose for at least seven days. We excluded those in the active phase of psychiatric illness, such as psychotic symptoms, who could not comprehend the assessment process instructions. Also excluded were the patients with signs of intoxication (Slurred speech, difficulty maintaining their balance, Slowed reaction, and aggressiveness. Those with a known history of severe cardiovascular condition since childhood, known congenital heart disease, or on treatment for cardiovascular conditions before opioid use were also excluded.

### Study variables

#### Outcome variables

The outcome variable was defined as having one or more electrographic Abnormalities included QTc prolongation in milliseconds (ms), sinus bradycardia, sinus tachycardia, left atrial enlargement, right atrial enlargement, and left ventricular hypertrophy. At first, it was dichotomized (Yes=presence of ECG abnormality /No= absence of ECG abnormality). The specific criteria for each ECG abnormality were referred from the the standard definition of terms is summarized in table 1 below.

**Table 1:**
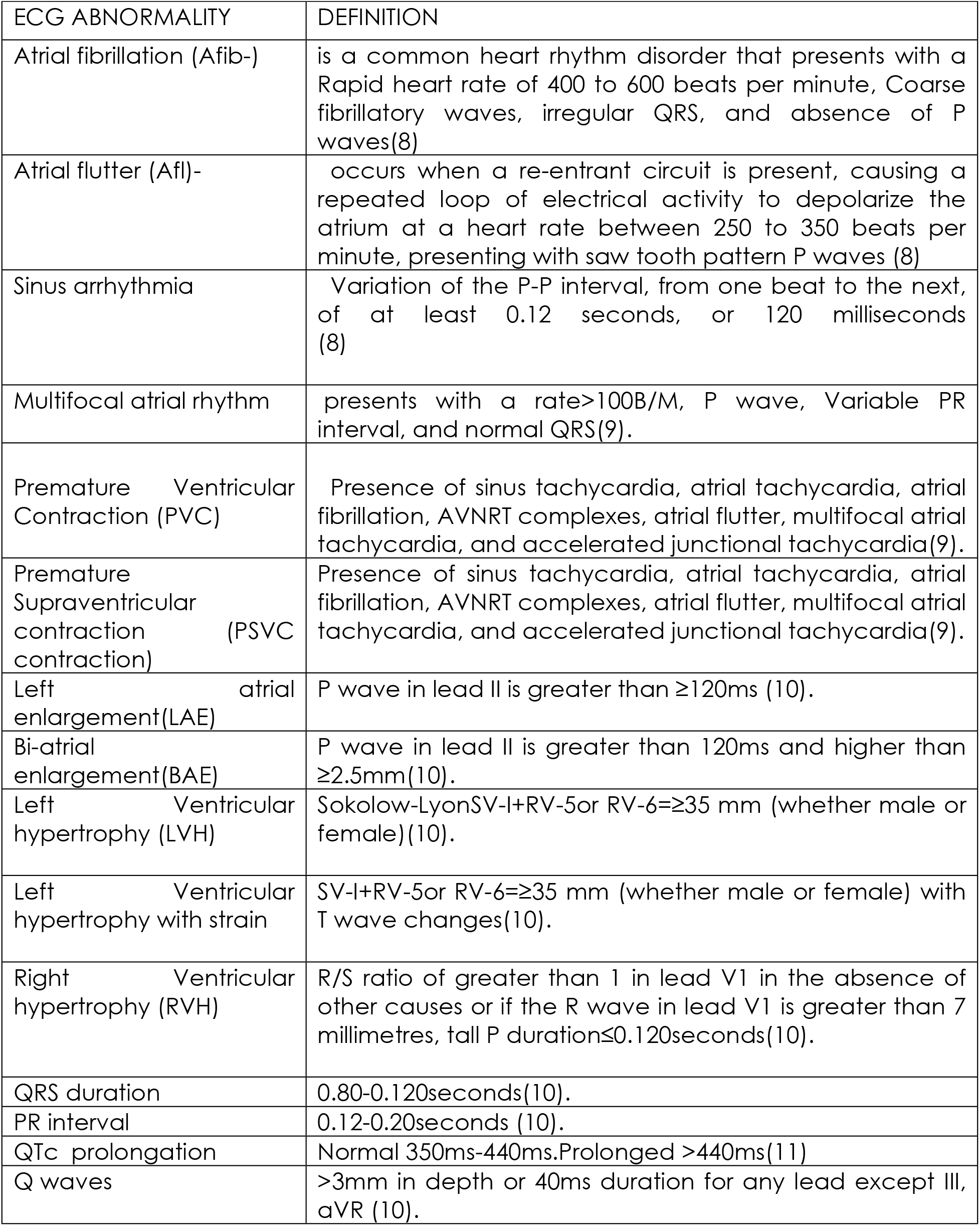
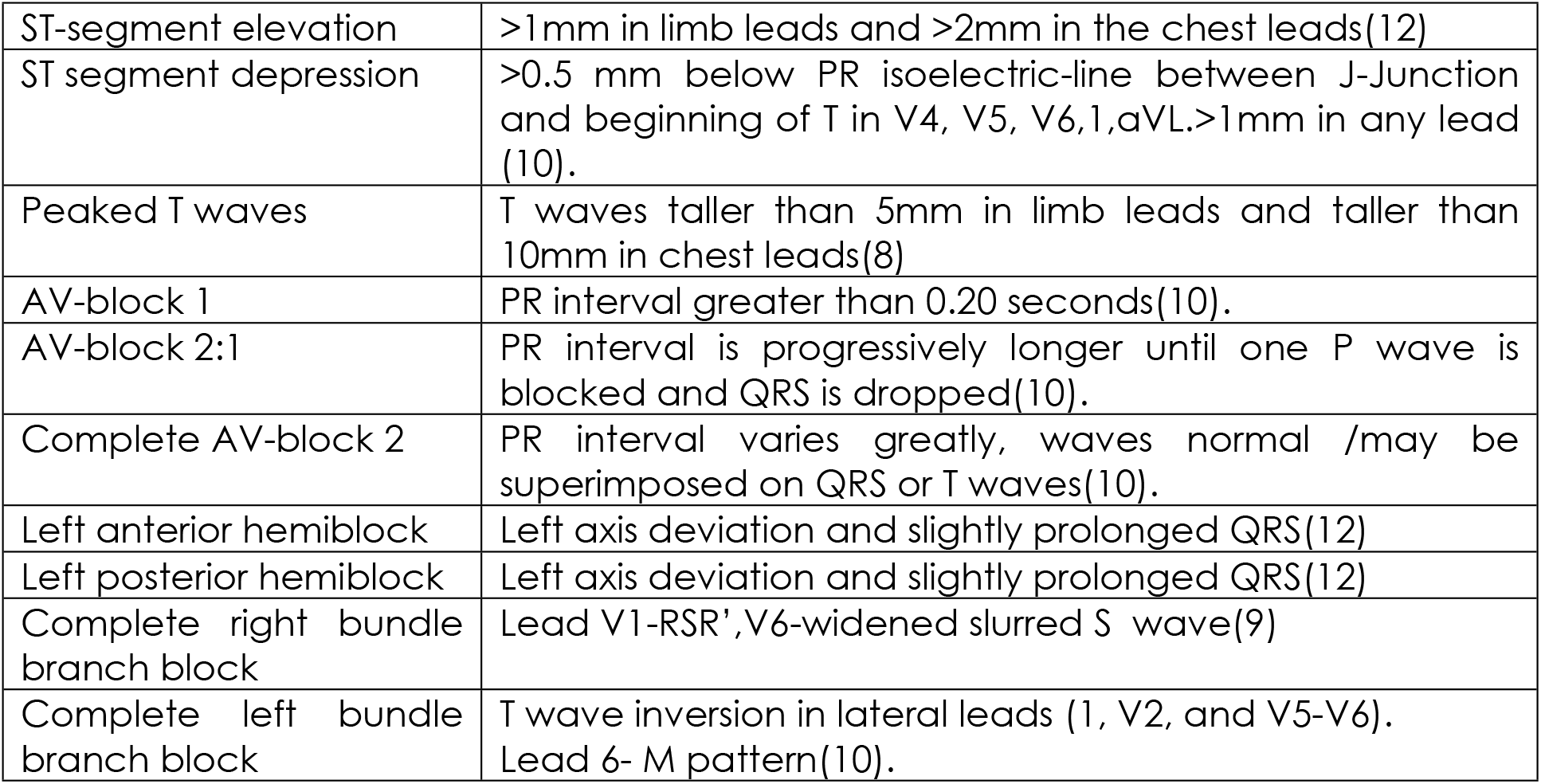
Summary of the standard definition for each ECG abnormality.

#### Independent variables

These included sociodemographic factors such as age (in years), sex, marital status, education, and employment status. Clinical factors included Hypertensive Heart disease (HTN), Diabetes Mellitus (DM), Anxiety and Depressive disorders, Hepatitis B and C viruses, and Body Mass Index (BMI). The substance use-related factors included smoking cannabis and cigarette, alcoholism, use of diazepam and cocaine within the past week, and concurrent substance use before methadone use. Opioid-related factors included the duration of opioid use, the duration of methadone treatment, and the current methadone dose in milligrams.

### Measurement of study variables

#### Electrocardiographic Abnormalities

A standard 12-lead ECG (BPL 9108) (SN H911180104201N000) machine manufactured by BPL medical technology in India was used. The participants had to wear a loose-fitting garment with buttons on the front, and women were asked to remove or loosen their brassiere. The participants lay supine, and electrode patches and leads were applied per the protocol. Upper limbs are placed between the elbow and shoulders.

Lower limb leads were placed a few inches above the ankle. Precordial leads were placed, V1, in the fourth intercostal space at the right border of the sternum. V2 is placed at the fourth intercostal space at the left border of the sternum. V3 is placed midway between V2 and V4. V4 is placed in the mid-clavicular line in the fifth intercostal space. V5 is placed in the anterior axillary line on the same horizontal level as V4 and V5. The participants were instructed to remain still while the ECG was obtained. Before recording, the display was checked for error messages, and a paper speed of 25mm per second and amplitude of 10mm/mV was used. The intervals (QT, QRS, and PR) were manually measured by hand using EKG calipers and rulers. PR interval, QRS duration, QT interval, QRS axis, Q, R, S, T-waves voltage, and ST-segment were measured in each lead. P-waves voltage was measured in lead V1 alone. Right atrial enlargement was defined as P wave voltage > 0.25 mV. Left atrial enlargement was defined as a biphasic P wave in V1. Left ventricular hypertrophy was calculated using Sokolow-Lyon voltage (sum of amplitude of the s waves in lead V1 and R waves in lead V 5 or V 6>3.5mV). Sinus bradycardia was considered when the heart rate was <60 B/M; sinus tachycardia was considered when the heart rate was>100 B/M. (8),(13).

The beginning of the QT interval was measured from the first deflection from the isoelectric line after the p wave. The end of the QT interval was defined at the point where the steepest tangent at the descending part of the T wave crosses the isoelectric line. The longest QT interval measured in the lead II, V2, or V5 from each ECG was used in further data analysis. QTc interval was calculated using Bazzet’s formula. Bazzett’s formula (QTc interval) = (QT interval) /√ (RR interval),(14,15). QTc interval >450ms for males and >470 for females was considered prolonged in this study (16) (17) (18).

#### Blood Pressure

An electronic blood pressure machine was used in measuring the blood pressure of all study participants who consented (HEM, Batch 7120, SN 1901006949, Omron Company, Kyoto, Japan). The participants were instructed not to smoke or take caffeine before taking the measurements. They sat in a chair with their back and arms supported at heart level. Blood pressure was measured after 5 minutes of rest; measurements were taken from the right arm. A cuff was placed approximately 2-5 cm above the ante-cubital crease. The machine was switched on, and then the start button was pressed to begin the inflation of the machine. Two readings were obtained, separated by five minutes intervals. The average value was recorded in the database.

#### Body Weight

A digital weighing machine was used for measuring body weight in kilograms (HL - 1, SN 1881041104024, Hesley Company, German). The scale was placed on a hard surface, and participants removed their shoes and were dressed in light clothing. The weight was recorded to the nearest 0.5 kg.

#### Height

A stadiometer scale was used for measuring height in meters (SECA, batch 217, Birmingham, United Kingdom). The participants removed their shoes and stood with their backs against the stadiometer. The participants were instructed to stand still with their heads at level with a stadiometer. The movable piece of stadiometer was fitted above the head of the participant. The height was recorded to the nearest meter.

#### Body Mass Index

The body mass index (BMI) was calculated as weight in kg divided by height in meter squares (kg/m^2^). Table 2 presents the categorization of BMI.

**Table 2:**
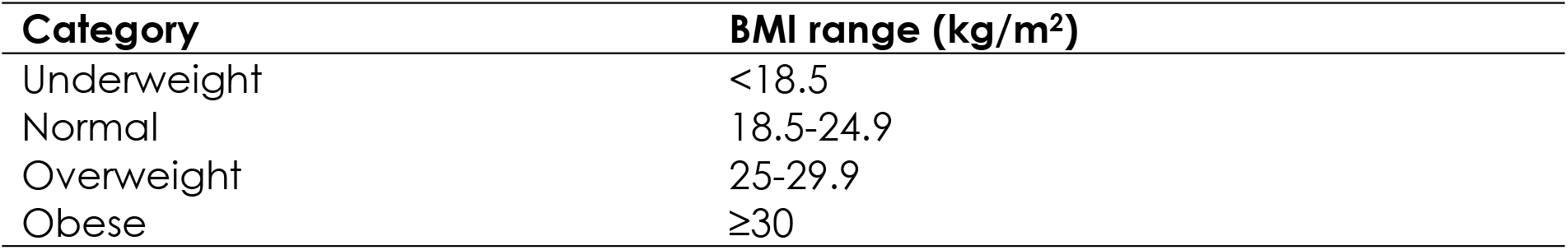
BMI classification (WHO, 2006)

#### Depressive and Anxiety symptoms

After enrollment in the MAT, the participants are screened for anxiety and depression after seven days as a routine.

Depression was screened using the PHQ-9 tool, which has been validated in Tanzania with a sensitivity of 78% and specificity of 87%, a cut-off score of 9 with a good internal consistency Cronbach’s alpha 0.83 (α = 0.83) (19). The PHQ −9 scores of 5,10,15, and 20 represented mild, moderate, moderately severe, and severe depression, respectively. (19) Anxiety was screened using a Swahili version of the GAD 7 tool, which has been validated in Kenya with good internal consistency (α = 0.82) (20).

#### Data collection procedure

Data were collected by the Principal investigator (PI) and five research assistants, including three medical officers and two registered nurses. The research assistants were trained in data collection for four days, whereby detailed familiarization with the data collection form and instruments for measuring variables. A data collection form was used to collect the required data (Appendix II). The data collection form comprised four sections. Section I: sociodemographic characteristics, section II: Clinical characteristics, section III: behavioral factors, and section IV: ECG findings and abnormalities. Interaction with the participants during data collection was done in a separate room to maintain privacy. It took approximately 30-50 minutes to complete the assessment and interview of each participant.

Pre-testing of data collection tools was done before the commencement of the study, whereby a pilot study was done two weeks before the commencement of the study. It included 20 clients on MAT, the trained research assistants interviewed the participants using the data collection forms, and the research instruments were used for measurable variables. This helped in assessing the applicability of the tools and provided an estimate of a timeframe for the interviews to make the necessary adjustments. Two research assistants (medical doctors) with special training in measuring ECG were given the task of placing ECG electrodes with the supervision of the PI. The ECG results were interpreted by two independent cardiologists blinded to patient history and clinical presentation, and in case of discordant interpretation, a third was to be consulted.

#### Data management and analysis

Data analysis was done using SPSS version 24, and cross-checking for the missing data and other errors was done after running frequency tables and crosstabs. The prevalence of ECG abnormalities was analysed by using descriptive statistics in which the overall prevalence of ECG abnormalities was determined and presented as a percentage. Categorical variables were summarized in proportions and percentages, while continuous variables were summarized in mean ± standard deviation (SD) or Median and Interquartile range. Binary logistic regression was used to determine the factors associated with ECG abnormalities. All variables with a p-value of ≤ 0.2 on univariate logistic regression were adjusted for multivariable analysis, and a p-value reaching ≤ 0.05 was considered statistically significant after adjusting for confounders. It should be noted that some variables succumbed to quasi-complete separation on multivariable logistic regression, therefore, were omitted.

## RESULTS

### Sociodemographic and Clinical Characteristics of Study Participants

The study included 321 participants with a mean age of 35.44 ± 5.55 years. The majority, 308 (95.95%), were males, 197 (61.37%) had attained primary educational level, 249 (77.57%) were single or never married, and being unemployed comprised 259 (80.69%) of the participants.

Regarding serological results, 52 (16.20%), 16 (4.98%), and 11 (3.43%) were found to have positive results for HCV, HIV, and HBV, respectively. Cigarette smoking 241 (75.08%) and cannabis 48 (14.95%) were the predominant concurrent substances used.

Over one-third, 123 (38.32%) of the participants were on >150 mg of methadone dose; sixteen participants (5%) met the criteria for hypertension, while 154 (47.98%) and 125 (38.94%) had prehypertension as per systolic and diastolic blood pressure parameters, respectively.

### Prevalence of ECG Abnormalities among Adults with Opioid Use Disorder Attending at Itega Addiction Centre for Methadone Assisted Therapy in Dodoma region

The prevalence of participants with one or more ECG abnormality was 85 (26.47%). (Figure 1).

**Figure 1:**
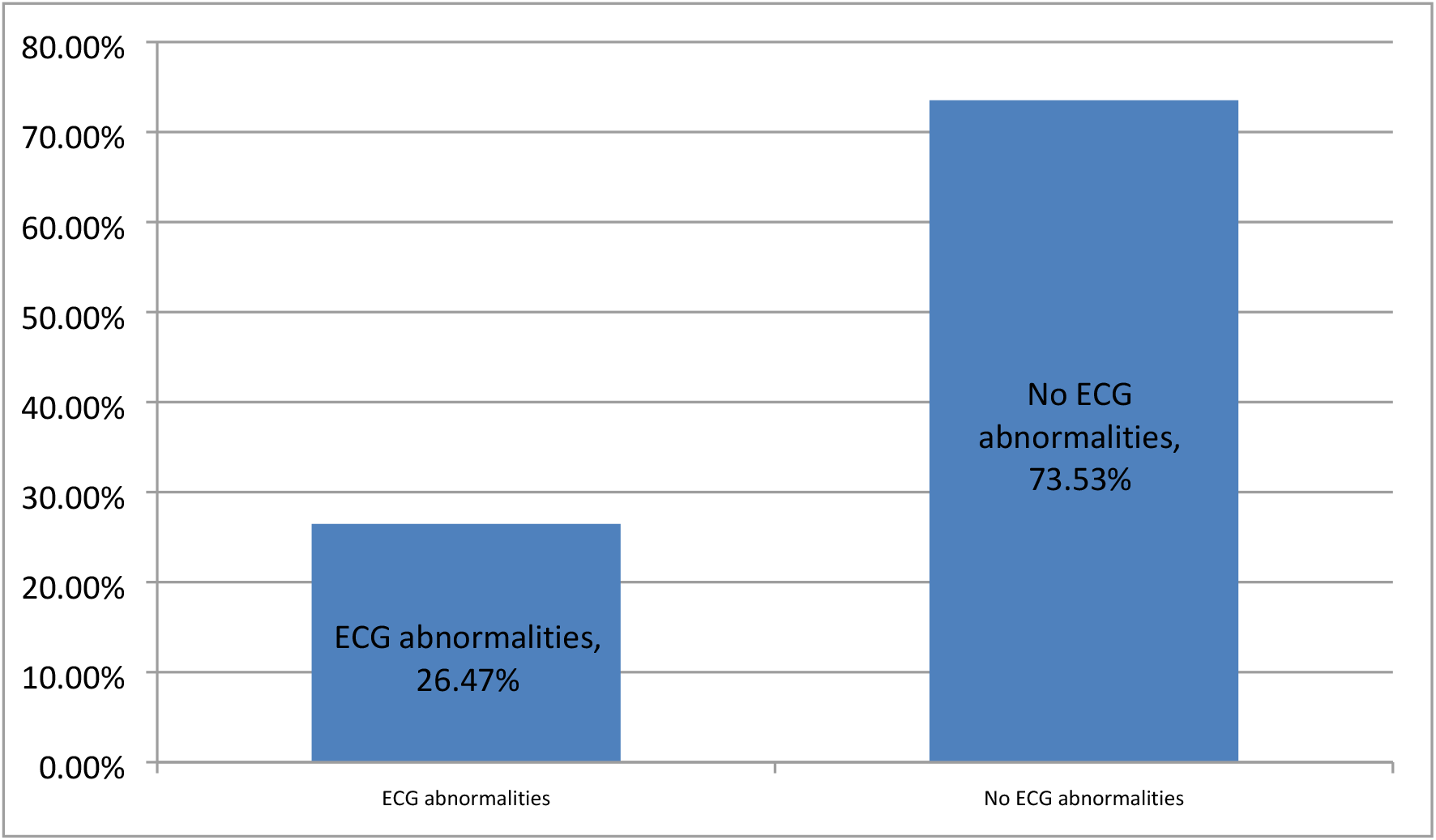
Prevalence of ECG Abnormalities among Adults with Opioid Use Disorder Attending at Itega Addiction Centre for Methadone Assisted Therapy in Dodoma region (N = 321)

### Identification of the ECG Abnormalities among Adults with Opioid Use Disorder Attending at Itega Addiction Centre for Methadone Assisted Therapy in Dodoma region

The most observed ECG abnormalities were sinus bradycardia 59 (18.4%) followed by prolonged QTc interval, which accounted for 27 (8.4%), while other ECG abnormalities had a frequency of 2(0.6%) or less, see Table 4.

**Table 3:**
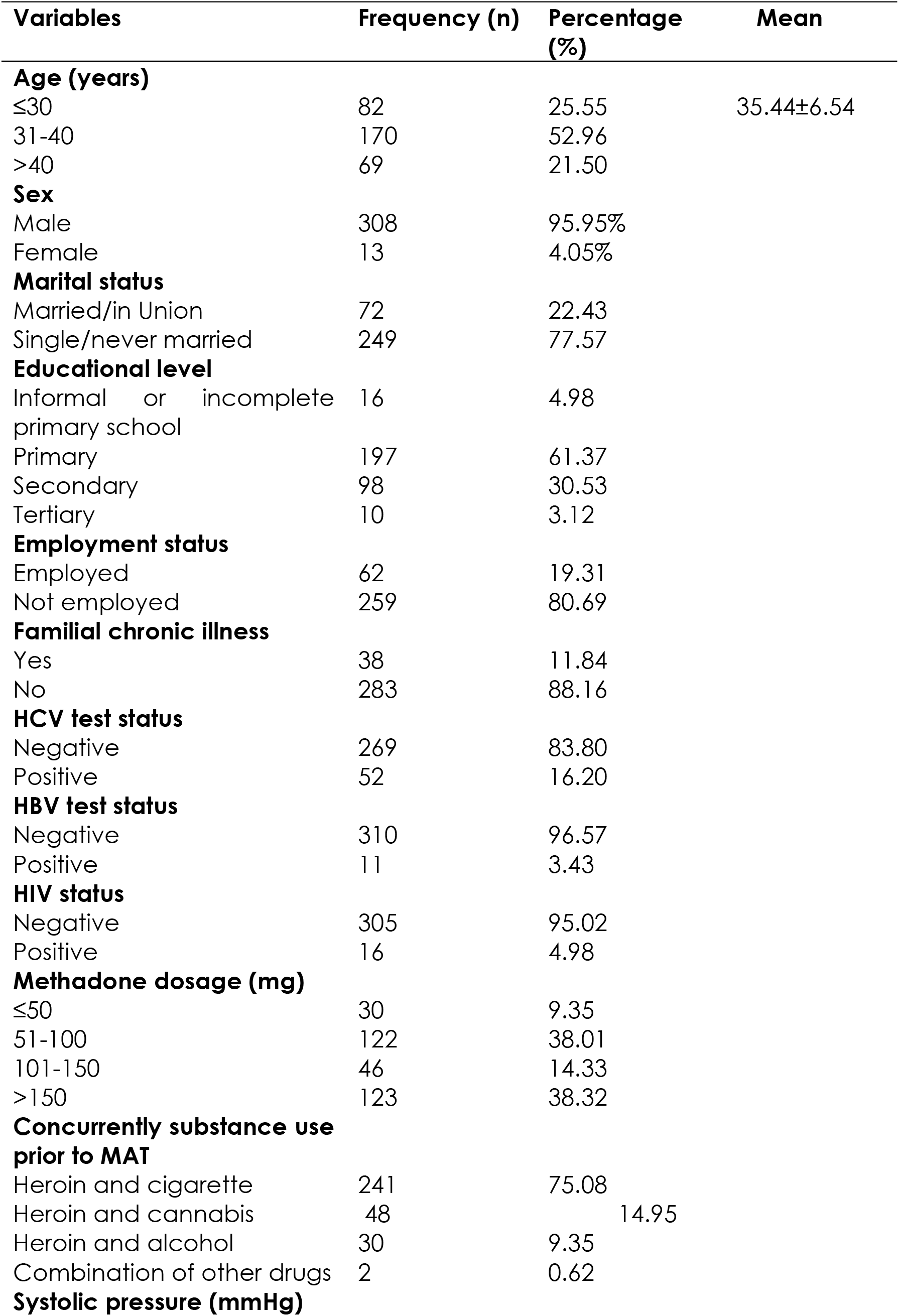
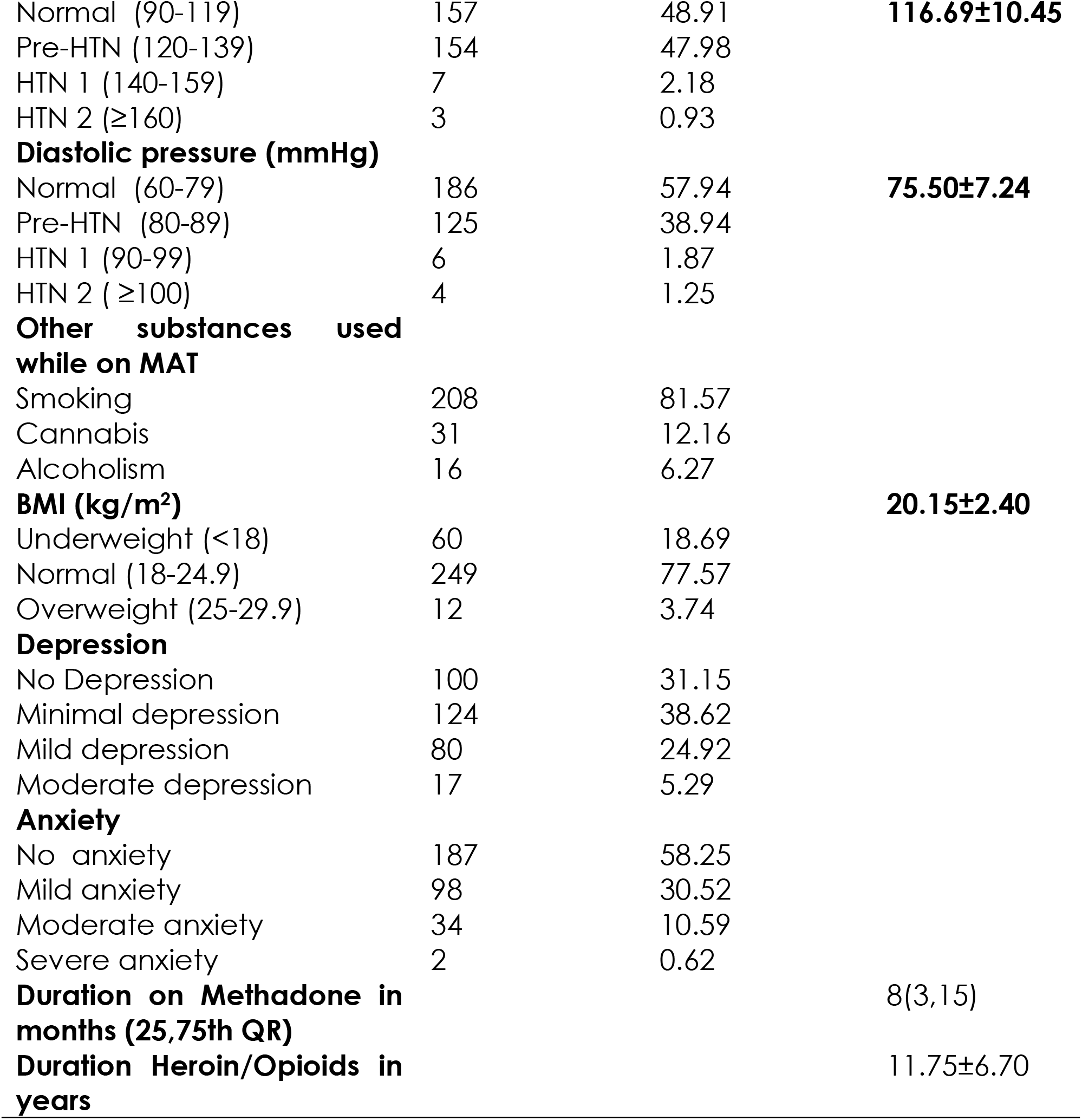
Sociodemographic and Clinical Characteristics of Adults Attending at Itega Addiction Centre for Methadone Assisted Therapy in Dodoma region (N = 321)

**Table 4:** Frequency Distribution of the ECG Abnormalities Among Adults with Opioid Use Disorder Attending at Itega Addiction Centre for Methadone Assisted Therapy in Dodoma region (N = 321)

| **ECG abnormalities\*** | **Frequency (n)** | **Percentage (%)** |
| --- | --- | --- |
| Sinus bradycardia | 59 | 18.4 |
| QTc prolongation | 27 | 8.4 |
| Sinus tachycardia | 2 | 0.6 |
| Left ventricular hypertrophy | 2 | 0.6 |
| Left atrial enlargement | 1 | 0.3 |
| Right atrial enlargement | 1 | 0.3 |

\*An individual may have more than one ECG abnormality

### Factors Associated with ECG Abnormalities

After adjusting for confounders, female sex (AOR = 0.05, 95% CI = 0.01-0.59, p = 0.017) and a month increase in the duration on MAT (AOR = 0.85, 95% CI = 0.74-0.96, p = 0.014) were significantly associated with decreased risk of ECG abnormalities.

Being underweight had a 1.24-fold increased risk for ECG abnormalities compared to normal weight, but the difference was not significant (AOR = 1.24, 95% CI = 0.65-2.37, p = 0.506). Also, having HBV and HIV infections significantly increases the risk of developing ECG abnormalities. (Table 5).

**Table 5:**
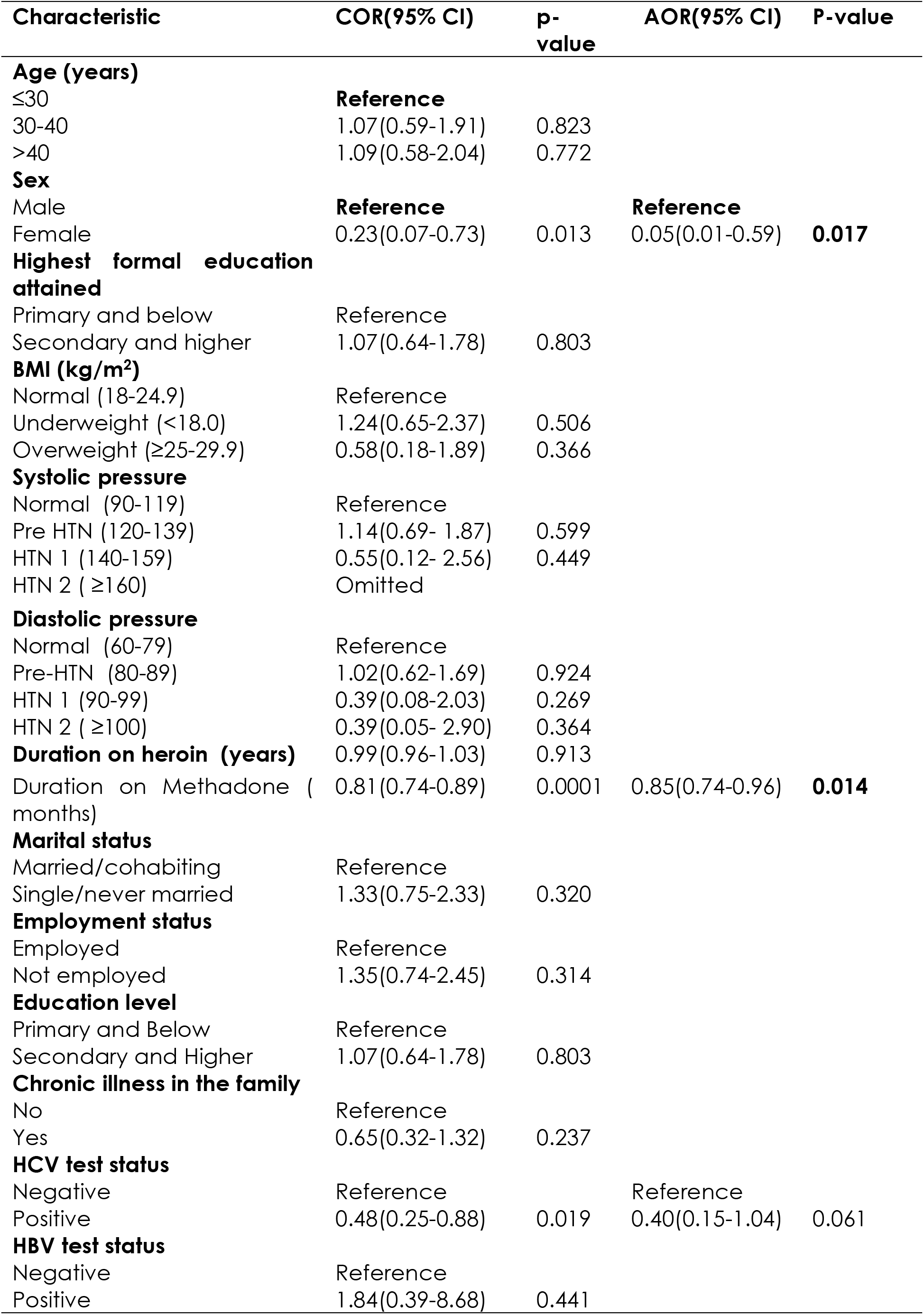
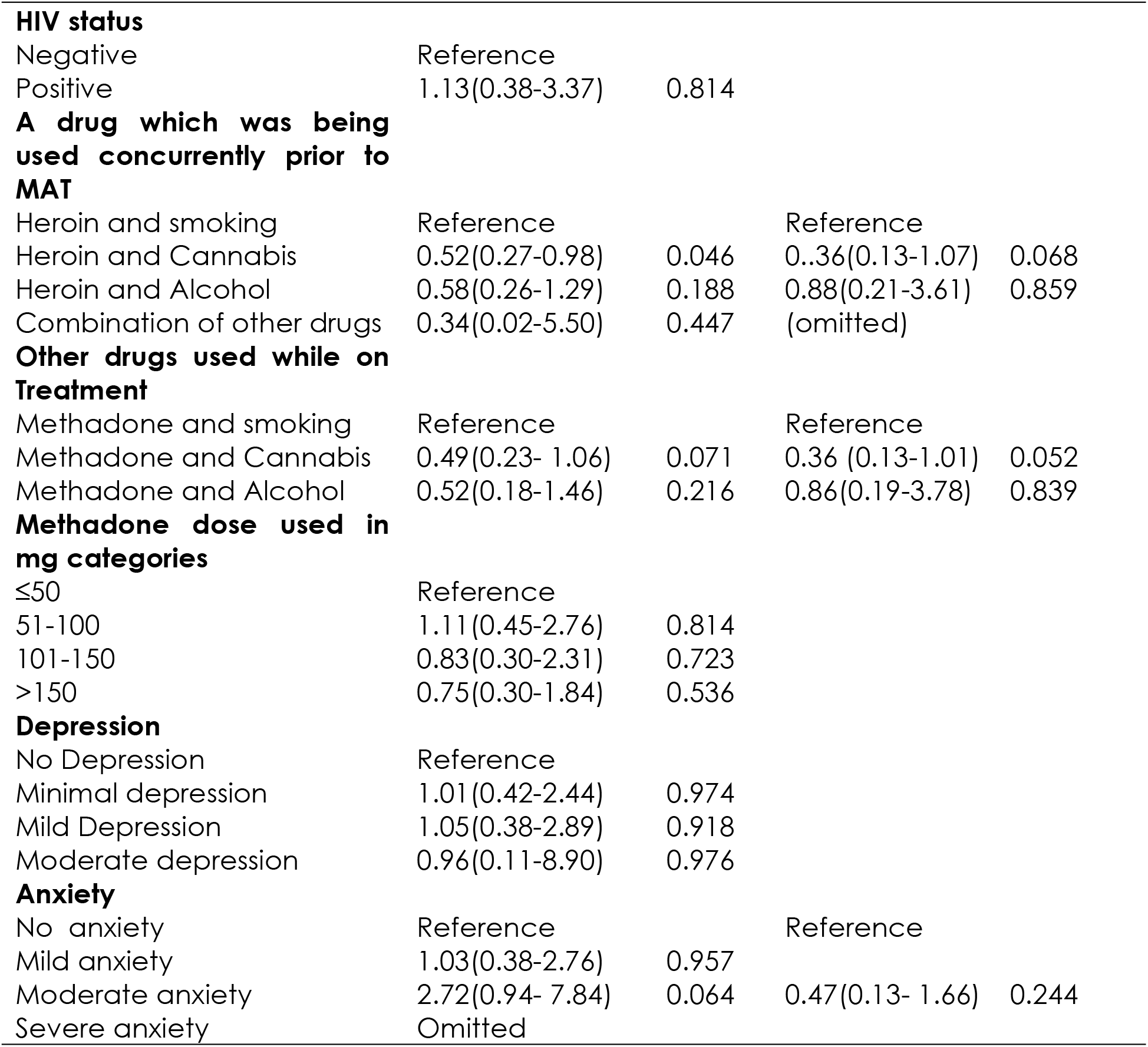
Factors Associated with ECG Abnormalities.

### Factors Associated with QTc Prolongation among Adults with Opioid Use Disorder Attending Itega Addiction Centre for Methadone Assisted Therapy in Dodoma region

Participants with moderate anxiety were almost four times the odds of having QTc prolongation compared to those without anxiety (AOR = 3.65, 95% CI = 1.18-11.26, p = 0.024). Being underweight significantly increased the risk of prolonged QTc interval compared to normal BMI (AOR = 2.90, 95% CI = 1.42-30.66, p = 0.027).

Although it was statistically significant, participants with pre-HTN by systolic pressure were almost twice more likely to be associated with QTc prolongation than those with normal systolic pressure (AOR = 1.72, 95% CI = 0.69-4.27, p = 0.236). Also, participants with mild anxiety had a 1.18-fold increased risk of getting QTc prolongation compared with those with no anxiety (AOR = 1.18, 95% CI = 0.42-3.31, p = 0.75). (Table 5).

### Factors Associated with Sinus Bradycardia

Under multivariable logistic regression, being underweight was significantly associated with the risk of sinus bradycardia compared to normal body weight (AOR = 5.19, 95% CI = 1.02-16.52, p = 0.048). Also, increased anxiety levels, such as mild compared to no anxiety (AOR = 0.19, 95% CI = 0.06-0.58, p = 0.004). Similarly, those who used cannabis while on methadone therapy were less likely to have sinus bradycardia compared to smoking cigarettes while on MAT (AOR = 0.23, 95% CI = 0.05-0.91, p = 0.037). A month increase in duration on MAT was also found to be significantly associated with sinus bradycardia (AOR = 0.74, 95% CI = 0.63-0.87, p = 0.001) (Table 6).

**Table 6:**
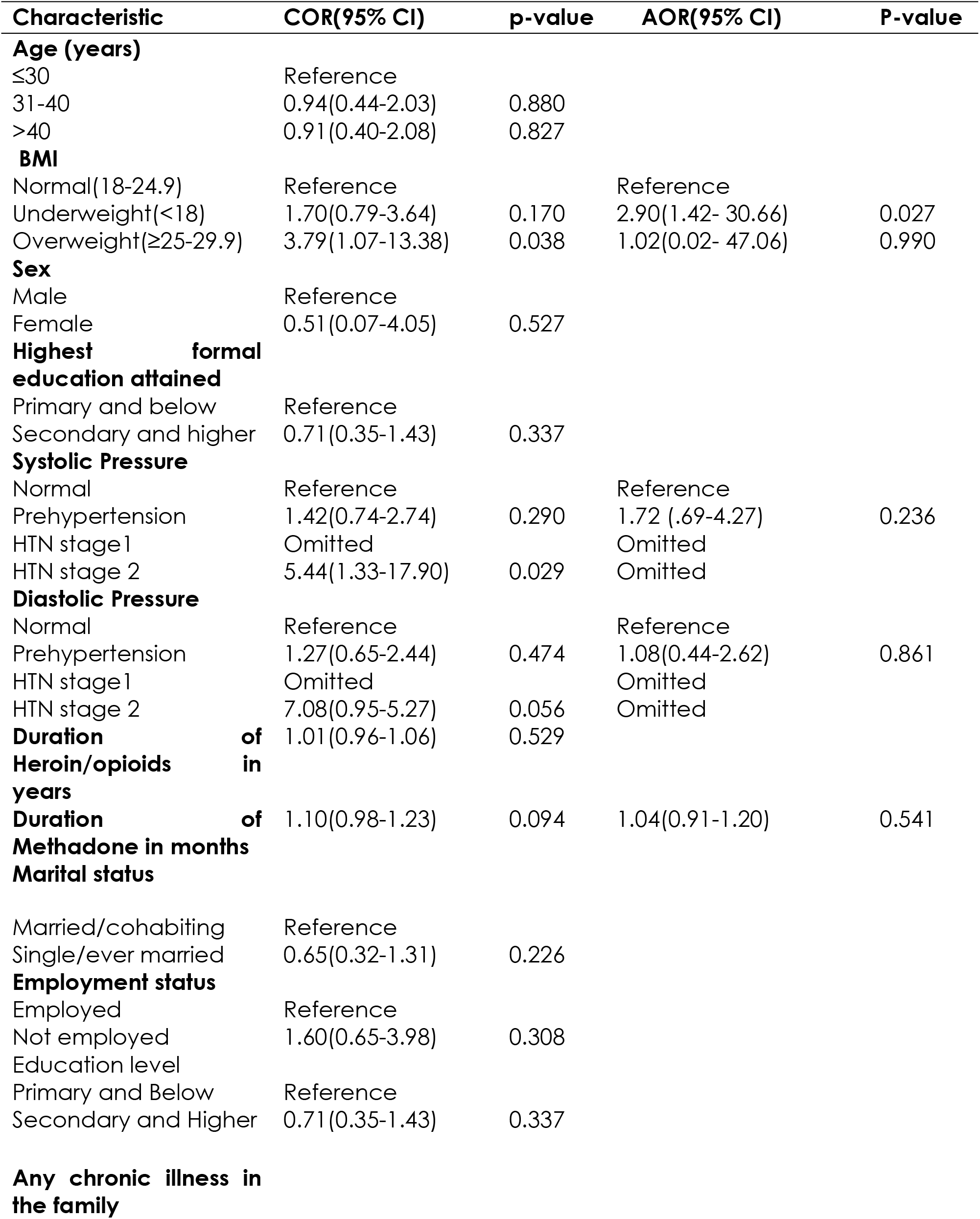
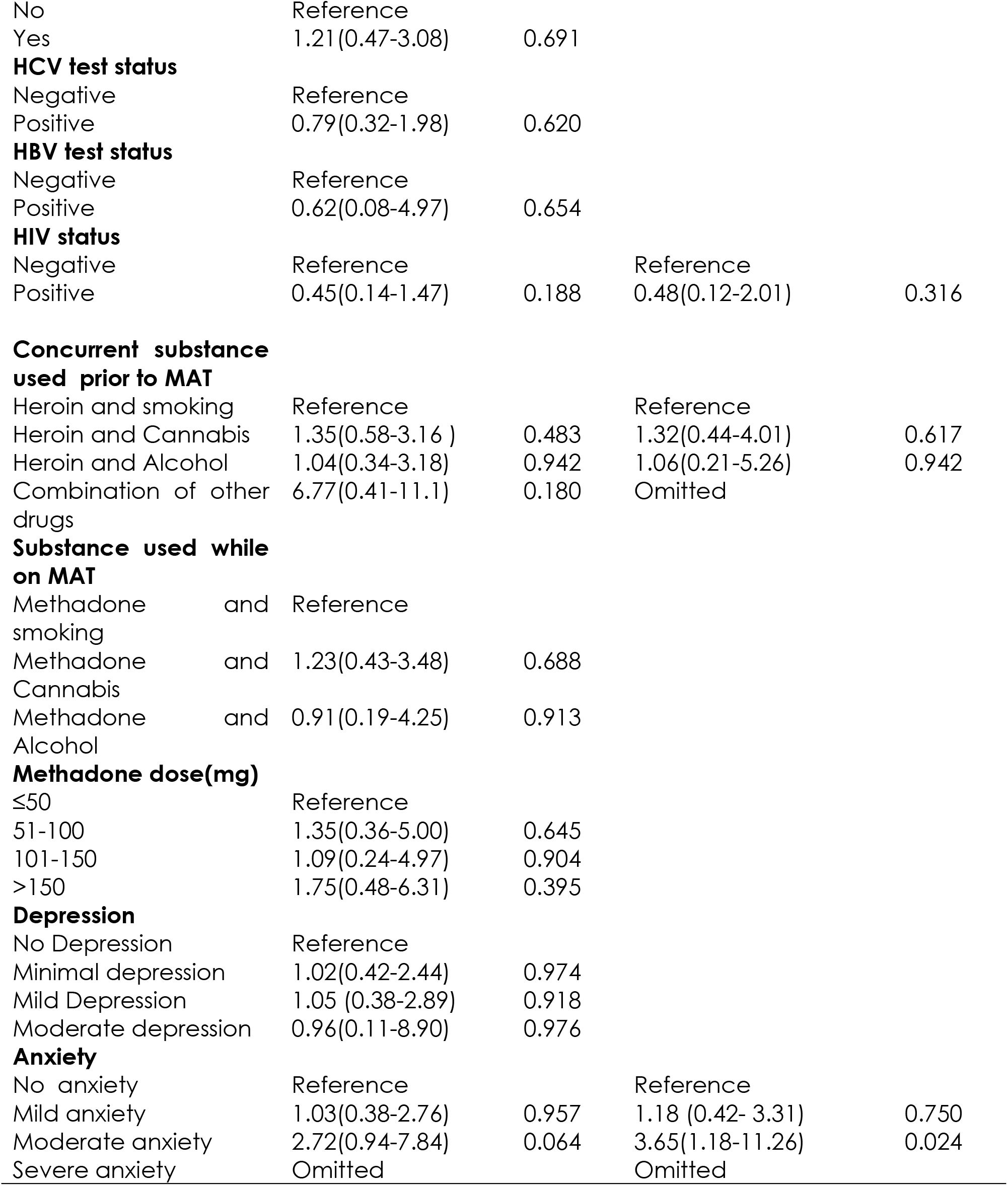
Factors Associated with QTc Prolongation Among Adults Attending at Itega Addiction Centre for Methadone Assisted Therapy in Dodoma region (N = 321)

**Table 1:**
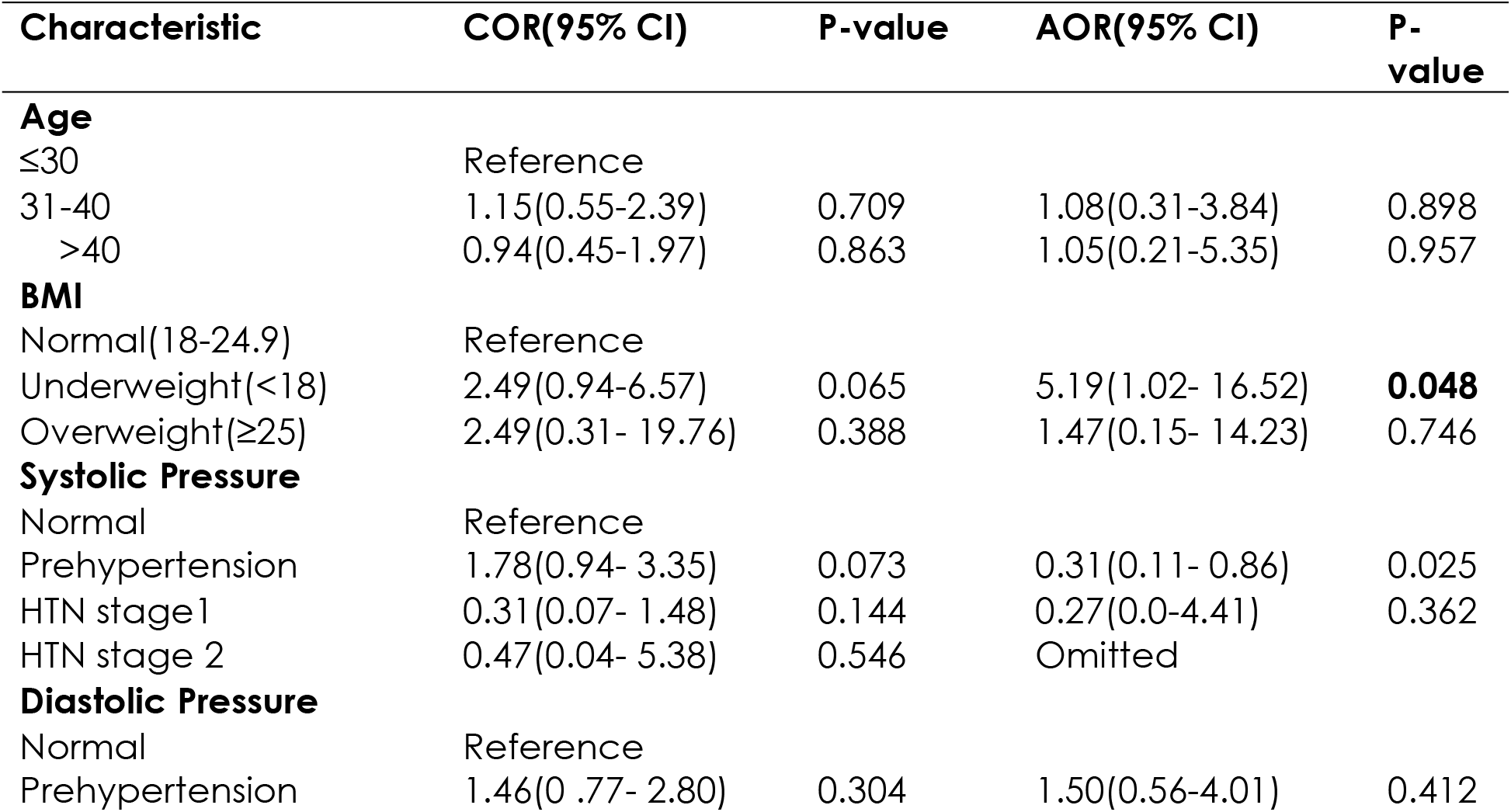
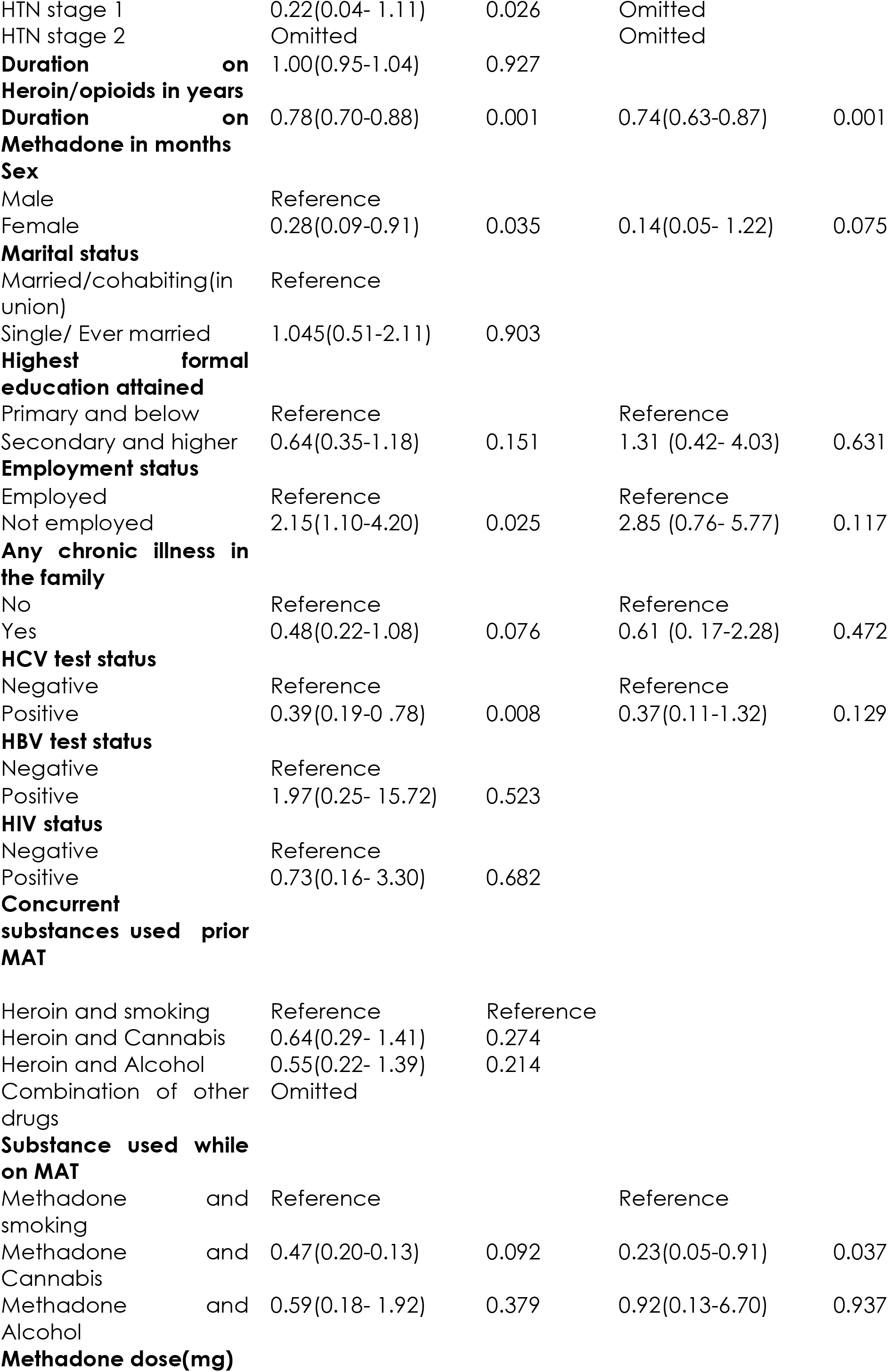
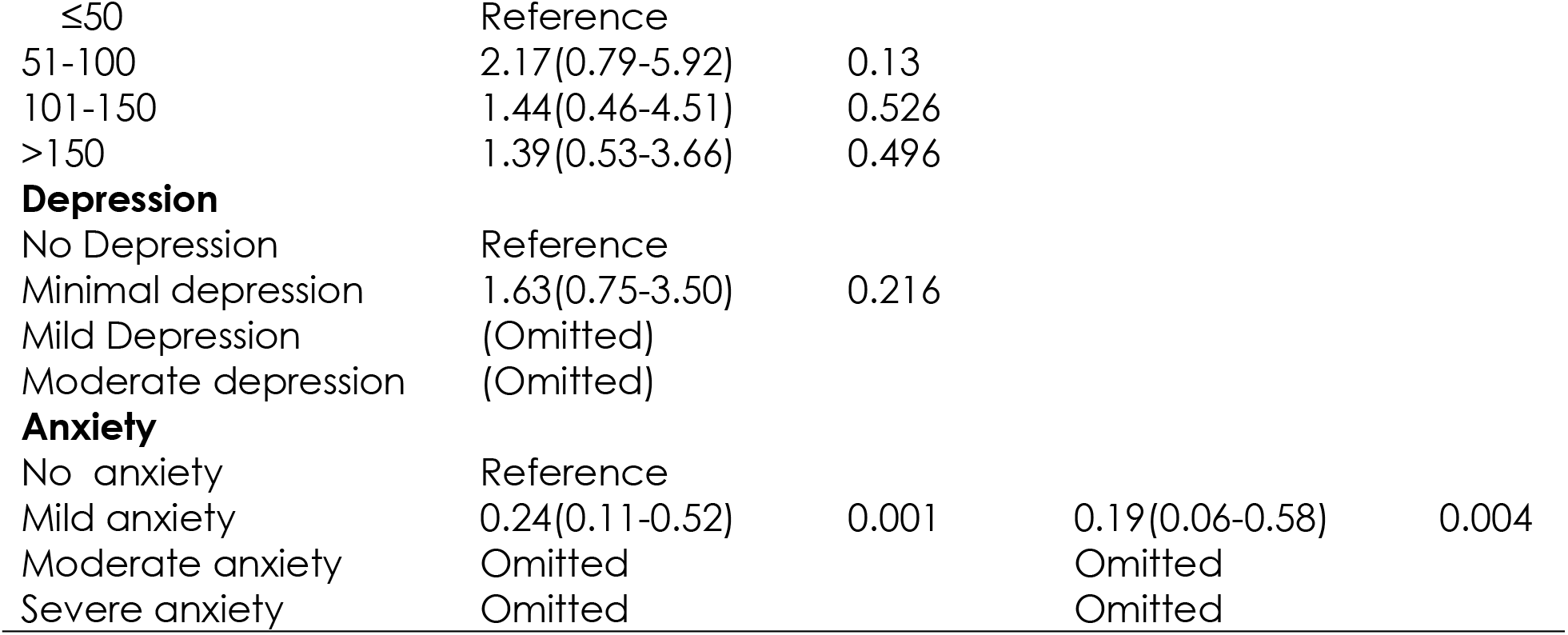
Factors Associated with Sinus Bradycardia Among Adults with Opioid Use Disorder Attending at Itega Addiction Centre for Methadone Assisted Therapy in Dodoma region.

## Discussion

The study observed a 24.47% prevalence of ECG abnormalities, with sinus bradycardia 59(18.4%) and QTc prolongation 27(8.4%) being the most prevalent of the ECG abnormalities.

The prevalence of ECG abnormalities varies globally, with higher rates of ECG abnormalities in MAT centers being reported. A 61% prevalence was reported in Austria(21). Martell et al, reported a prevalence of 48.3% among 118 patients with opioid use disorder on methadone treatment (22), a prevalence of 54% in the United States of America(23), and 59.3% in India(24). The variations in the study population may explain the diversity in prevalence. In contrast, this study had younger; a study in India included opioid users who were older and with comorbid non-cardiac medical conditions(24).

The role of aging as an independent risk of ECGs abnormalities, including greater QTc, has been demonstrated(22,25). Aging processes may affect the molecular determinants of the QT interval or alter the myocardium with increased myocardial fibrosis previously observed in the Multi-Ethnic Study of Atherosclerosis study(26). Aging is also associated with alterations in the amount of sympathetic and parasympathetic tone, that alters myocardial repolarization and the duration of the QTc (27). Despite the interaction with other factors, opioid drugs, in general, can uniquely affect the ECG and lead to the development of cardiac arrhythmias and QTc interval prolongation being the most significant side effect (28). The mechanism of QTc prolongation due to opioid use is explained by inhibiting the Human ether a go-go related gene(hERG)(29). Also, negative chronotropic effects via Ca^+^ channel antagonism and anticholinesterase effect explain the mechanisms of opioid-induced cardiac arrhythmias(30).

Similar to our study, QTc prolongation and sinus bradycardia are also observed to be the most common abnormalities among people with opioid addiction (22, (23). In contrast, few others report ST abnormalities to be the most common ECG abnormalities (21). Other less common ECG abnormalities with a smaller proportion include Brugada syndrome, sinus tachycardia, and ST changes (31). While the study may suggest a high prevalence of sinus bradycardia based on the criteria of a pulse rate of < 60 beats a minute, this may be a normal observation in healthy young adults and older adults above 65 years. Healthy young adults have increased vagal tone, which could be the cause of sinus bradycardia at rest (32). Nevertheless, the over-representation of sinus bradycardia could also be attributed to heroin acting centrally on the vasomotor center to increase parasympathetic and reduce sympathetic activity (33) and also slowing down the sinus node, which is responsible for the regulation of cardiac electrical activity (34).

Methadone being an opioid, chronic use could be linked to cardiotoxicity related to several ECG abnormalities, including QT dispersion, torsades-de pointes, pathological U waves, ventricular bigeminy, Takotsubo syndrome, Brugada-like syndrome and coronary artery diseases(36). Several factors are also associated with ECG abnormalities; previous studies have linked higher dosages of methadone to QTc prolongation(30). A study in Iran showed a 54.6% prevalence of QTc prolongation, which is attributed to a higher dosage of methadone compared to the dosage of < 150mg provided for the majority of our participants (35). Interestingly, a month-unit increase in the duration of methadone in two years of use was associated with fewer ECG abnormalities in our study; participants who are on longer duration of methadone use in years are observed to have a higher prevalence of ECG abnormalities(23). While the observation may suggest a cardiovascular benefit of methadone treatment, at least in the early phase of MAT, prolonged use of methadone may have been linked to ECG abnormalities.

Our study showed that being male had a positive association with ECG abnormalities. Generally, males are observed to have higher cardiovascular risk at a relatively younger age (37) compared to female counterparts exposed to cardioprotective endogenous estrogen during the fertile period. However, the benefit is lost post-menopausal (38).

Increased anxiety levels were associated with QTc prolongation but inversely associated with sinus bradycardia. Although Piccirillo et al showed that anxiety, even at moderate levels, was associated with QTc prolongation (39), other studies found no association (40). Anxiety is thought to disturb the cardiac autonomic balance and is related to an increased risk of cardiac arrhythmia (41) and also mediates increase in QTc interval variability via adrenergic mechanisms(42), atrial fibrillation(43), and coronary heart diseases (44). The physiological mechanism of anxiety in causing cardiovascular diseases includes inflammation, endothelial dysfunction, platelet dysfunction, and autonomic dysfunction(45). Anxiety state can also trigger an increase in neurotransmitters such as dopamine and epinephrine which cause an increased heart rate(46) although sinus bradycardia has been reported in anxiety disorders(47).

The study also showed that being underweight was significantly associated with ECG abnormalities. Being Underweight has been linked to QTc prolongation, sinus bradycardia and ventricular arrhythmias (48,49,50,53,54) which could be associated with micronutrients, macronutrients deficiencies, and electrolytes imbalances(49,51,52).

A possible cause of ECG abnormalities among underweight is the loss of cardiac muscle mass associated with cardiac conduction defects(55,56). Sinus bradycardia is a common phenomenon in the underweight population as the body’s parasympathetic nervous systems attempt to conserve energy(49,57), as the negative energy balance has a role in the variation of the autonomic nervous system with weight loss(58).

Another factor that predicted sinus bradycardia was cigarette smoking, an observation that could be attributed to nicotine which is the main constituent of cigarettes and is a potent blocker of the cardiac A-Type K^+^ channels, which cause-effect on cardiac physiology(59). While nicotine is thought to stimulate catecholamine release by activating nicotine acetylcholine receptors in peripheral postganglionic sympathetic nerve ending and adrenal medulla leading to cardiac tachycardia(60,61), a paradoxical effect of nicotine through its effect on A-type K^+^ leading to bradycardia has also been observed(59).

The study had several limitations; first, the cross-sectional study design could not establish a causal relationship between the predictor and outcome variable. Second, since we only used a single 12-lead ECG to detect ECG abnormalities, diurnal variations of QTc measurements cannot be established. Third, a few potential confounders, including the serum electrolytes, urine drug screen, and serum methadone levels, were not measured and could not be accounted for. Lastly, the disproportionate overrepresentation of male study participants may mask the actual gender difference associated with ECG abnormalities.

### Conclusion and recommendation

The study shows significant proportion of people with opioid addiction is at increased risk electrocardiographic abnormalities. While, ECG abnormalities pose the risk of cardiovascular related morbidity and mortality, their impact could potentially be prevented by integrating early cardiovascular care in routine MAT clinics. Furthermore, studies employing different design including prospective cohort would offer more information causal relationship and platform to design evidence based interventions.

### Ethical issues

Ethical approval was obtained from the University of Dodoma with reference number “**NO.UDOM/RP/68/Vo1.IV/10**”. Permission to conduct the study was obtained from Itega addiction Centre. Written informed consent forms were provided for those who could read and write; otherwise witnessed verbal informed consent was to be used as an alternative. No minor was included in the study, if a participant could not consent due to medical or any other reason, a custodian or close relative provided the consent. The participants were free to withdraw from the study at any point in time with affecting their daily routine care, and those found with ECG abnormalities were scheduled for cardiology clinic for further investigation and follow up.

## Data Availability

Data will be shared as supplementary file under request

